# Sociodemographic Predictors of Telehealth Physical Therapy Utilization in the United States: Analysis of the 2023 MEPS Data

**DOI:** 10.64898/2025.12.03.25341606

**Authors:** Elton J Tidwell

## Abstract

**Background:** Telehealth physical therapy (PT) expanded rapidly during the COVID-19 pandemic, but patterns of utilization across populations remain poorly understood.

**Objective:** To examine sociodemographic predictors of telehealth PT use in a nationally representative U.S. sample.

**Methods:** Data from the 2023 Medical Expenditure Panel Survey (MEPS) were analyzed. Adults with physical therapy visits were included. Weighted descriptive statistics and logistic regression models were used to estimate associations between TelePT use and age, education, sex, race/ethnicity, and income.

**Results:** Among 13,800 adults with PT visits, 1,590 (11.5%) used TelePT. Weighted analyses showed TelePT users were more likely to be female (62% vs 53%), White (80% vs 76%), and middle/high income (81% vs 76%). Logistic regression indicated higher odds of TelePT use with increased education (OR 1.17, 95% CI 1.15–1.19), female sex (OR 1.34, 95% CI 1.21–1.51), and White race (OR 1.24, 95% CI 1.08–1.41). Age (OR 0.98, 95% CI 0.98–0.98) was associated with reduced odds. Income was not statistically significant after adjustment.

**Conclusions:** TelePT adoption is uneven across sociodemographic groups, with higher use among more educated, female, and White patients. These findings help identify those most likely to adopt TelePT services and these disparities highlight the need for targeted strategies to ensure equitable access to telehealth physical therapy.

## Introduction

Telehealth has transformed the delivery of healthcare services by expanding access and reducing barriers related to geography and mobility [1,2]. During the COVID-19 pandemic, policy shifts and rapid technological adoption led to a dramatic increase in the use of telehealth for physical therapy (PT) and rehabilitation [3,4]. Telehealth PT has demonstrated effectiveness for musculoskeletal conditions, postoperative rehabilitation, and chronic disease management, with outcomes comparable to in-person care [5,6].

Despite its promise, telehealth adoption has not been uniform across patient populations. Prior research suggests that sociodemographic factors such as age, race, education, and socioeconomic status influence telehealth utilization [7,8]. Younger adults and those with higher digital literacy tend to adopt telehealth more readily, while older adults face barriers related to technology access and usability [9]. Disparities in access among racial/ethnic groups and across income strata remain a concern [10].

Understanding which populations are more likely to access PT through telehealth is critical for designing equitable and sustainable service delivery models. To date, few studies have examined sociodemographic predictors of telehealth PT utilization in the United States using nationally representative data. This study uses the 2023 MEPS to identify demographic and socioeconomic factors associated with telehealth PT use.

## Methods

### Data Source

We used data from the 2023 Medical Expenditure Panel Survey (MEPS), conducted by the Agency for Healthcare Research and Quality (AHRQ). MEPS is a nationally representative survey of the U.S. civilian non-institutionalized population and is widely used for health services research [13].

### Study Sample

The analytic sample included adults (≥18 years) with at least one PT/OT visit reported in 2023. After merging and applying survey weights, the final sample comprised 13,409 adults.

### Measures

Outcome: Telehealth use for PT/OT visits (TELEHEALTHFLAG variable, 1 = telehealth, 0 = in-person).

Predictors: Age (years), sex (male vs female), race (collapsed to White vs Non-White), education (years), poverty status (Low vs Middle/High).

Survey design: Person-level weights (PERWT23F), variance strata (VARSTR), and primary sampling unit (VARPSU) were used.

### Statistical Analysis

Weighted descriptive statistics compared TelePT vs non-TelePT users. Group differences were tested with unweighted t-tests (continuous variables) and chi-square tests (categorical). Logistic regression models estimated odds of TelePT use, adjusting for all predictors simultaneously. Odds ratios (OR) with 95% confidence intervals (CI) were reported. Robust standard errors accounted for survey design.

## Results

### Sample Characteristics

Final analytic sample: 13,800 adults (weighted N ≈ U.S. adult PT population).

- TelePT users: 1,590 (11.5%).
- Weighted mean age: 42.6 years for TelePT vs 42.1 years for non-TelePT (p = 0.98).
- Education: higher among TelePT users (13.7 vs 11.0 years, p < 0.001).
- Female: more common in TelePT users (62% vs 53%, p < 0.001).
- White race: more common in TelePT users (80% vs 76%, p = 0.0001).
- Middle/High income: more common in TelePT users (81% vs 76%, p < 0.001).

**Table 1.** Weighted Sample Characteristics.

| Characteristic | No TelePT (weighted) | TelePT (weighted) | Unweighted N | Unweighted p-value |
| --- | --- | --- | --- | --- |
| Sample size | – | – | 12,403 / 1,590 | – |
| Age, years (mean ± SD) | 42.1 ± 24.9 | 42.6 ± 19.3 | – | 0.9803 |
| Education, years (mean ± SD) | 11.0 ± 5.8 | 13.7 ± 3.9 | – | <0.001 |
| Female, % | 53.1% | 61.7% | – | <0.001 |
| White, % | 76.4% | 79.6% | – | 0.0001 |
| Middle/High income, % | 75.7% | 81.0% | – | <0.001 |

**Notes:**
- Weighted estimates use person-level survey weights (PERWT23F). - P-values are from unweighted t-tests (for continuous variables) or chi-square tests (for categorical variables).

### Regression Analysis

#### Logistic regression (weighted, robust SEs)

- Age: OR 0.98 (95% CI 0.98–0.98, p < 0.001).
- Education: OR 1.17 (95% CI 1.15–1.19, p < 0.001).
- Female vs Male: OR 1.34 (95% CI 1.21–1.51, p < 0.001).
- White vs Non-White: OR 1.24 (95% CI 1.08–1.41, p = 0.002).
- Income (middle/high vs low/near): OR 0.98 (95% CI 0.86–1.13, p = 0.81).

**Table 2.** Logistic Regression Results.

|  | Predictor | OR | CI_low | CI_high | p-value |
| --- | --- | --- | --- | --- | --- |
| Age (years) | Age (years) | 0.98 | 0.98 | 0.98 | 0.00 |
| Years of education | Years of education | 1.17 | 1.16 | 1.19 | 0.00 |
| Sex: Female vs Male | Sex: Female vs Male | 1.35 | 1.21 | 1.51 | 0.00 |
| Income: Middle/High vs Low/Near | Income: Middle/High vs Low/Near | 0.98 | 0.86 | 1.13 | 0.81 |
| Race: White vs Non-White | Race: White vs Non-White | 1.24 | 1.08 | 1.41 | 0.00 |

**Figure 1.**
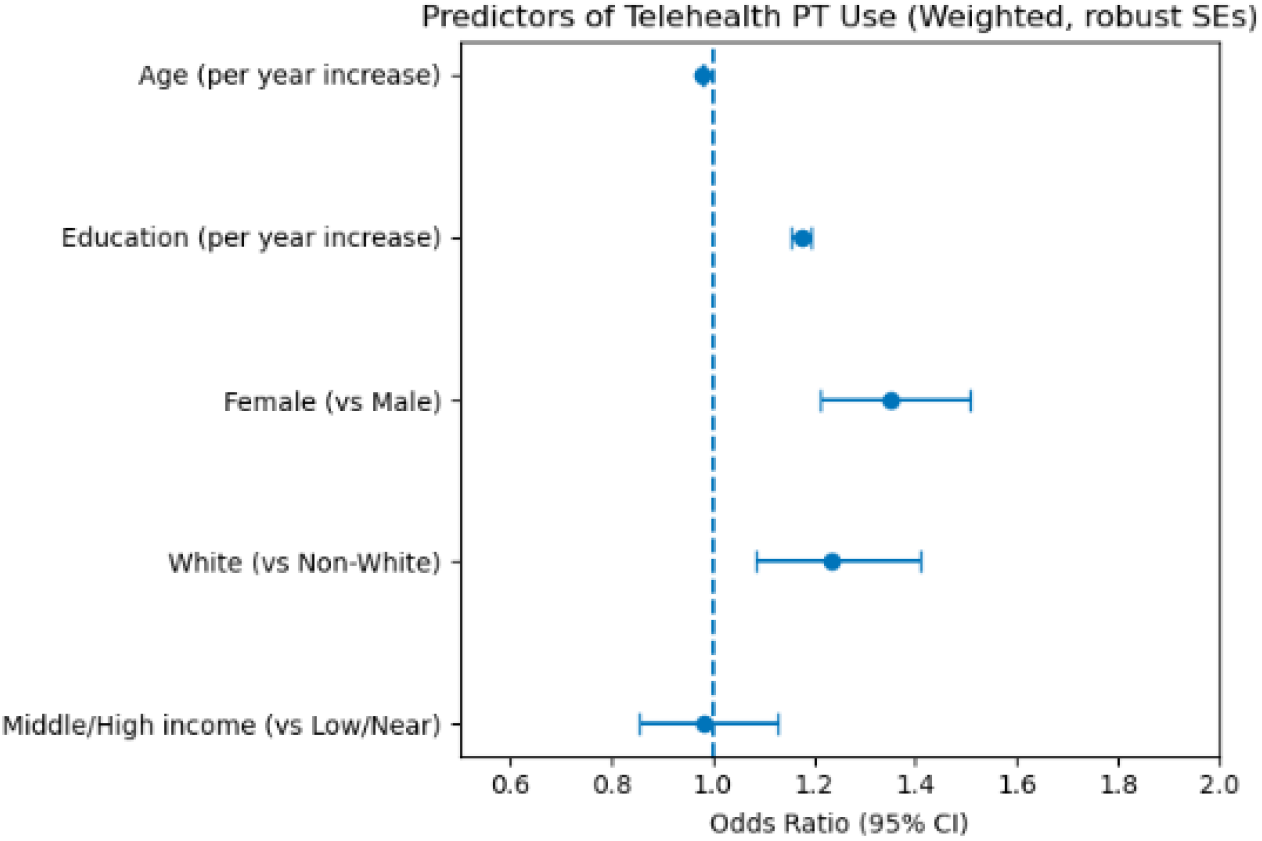
Forest Plot of Odds Ratios

## Discussion

In this nationally representative analysis, TelePT use was more common among younger, more educated, female, and White adults. These findings mirror broader disparities in telehealth adoption [7–12] and suggest that while TelePT can reduce geographic barriers, it may inadvertently widen gaps across sociodemographic groups.

Notably, income was not a significant predictor after adjustment, which may reflect the influence of insurance coverage and targeted telehealth policies during the pandemic. Education remained a strong predictor, highlighting the role of digital literacy in accessing virtual care.

These findings underscore the need for targeted interventions—such as patient education, simplified telehealth platforms, and outreach in underserved communities—to ensure equitable TelePT access.

## Conclusion

Telehealth physical therapy use is rising but unevenly distributed. More educated, female, and White patients are more likely to use TelePT, while older adults remain less likely to adopt. This information can be used to target those most likely to accept telehealth services. Future efforts should focus on reducing digital literacy barriers and ensuring equitable access across diverse populations.

## Data Availability

All data analyzed in this study are derived from the publicly available Medical Expenditure Panel Survey (MEPS) datasets, accessible through the Agency for Healthcare Research and Quality (AHRQ) website:
https://meps.ahrq.gov/mepsweb/data_stats/download_data_files.jsp
No proprietary data were used, and all analyses are fully reproducible using these open data sources.

